# Long COVID is associated with extensive *in-vivo* neuroinflammation on [^18^F]DPA-714 PET

**DOI:** 10.1101/2022.06.02.22275916

**Authors:** Denise Visser, Sandeep S.V. Golla, Sander C.J. Verfaillie, Emma M. Coomans, Roos M. Rikken, Elsmarieke M. van de Giessen, Marijke E. den Hollander, Anouk Verveen, Maqsood Yaqub, Frederik Barkhof, Janneke Horn, Bart Koopman, Patrick Schober, Dook W. Koch, Robert C. Schuit, Albert D. Windhorst, Michael Kassiou, Ronald Boellaard, Michele van Vugt, Hans Knoop, Nelleke Tolboom, Bart N.M. van Berckel

## Abstract

A significant number of COVID-19 patients develop ‘long COVID’, a condition defined by long-lasting debilitating, often neurological, symptoms. The pathophysiology of long COVID is unknown. Here we present *in-vivo* evidence of widespread neuroinflammation in long COVID, using a quantitative assessment, [^18^F]DPA-714 PET, in two long COVID patients. We reanalyzed historical data from three matched healthy control subjects, for comparison purposes. Both patients with long COVID had widespread increases in [^18^F]DPA-714 binding throughout the brain. Quantitative measures of binding (BP_ND_ values) were increased on average by 121% and 76%, respectively. This implicates profound neuroinflammation in the pathophysiology of long COVID.

## Introduction

Approximately 36-53% of COVID-19 patients develop long-lasting, chronic complaints, a condition known as post-acute sequelae of SARS-CoV-2 infection (PASC) or ‘long COVID’ (1-4). Although in the acute phase of infection, COVID seems to primarily affect the respiratory system, its long-term effects are mainly neurological. The most frequently reported symptoms are fatigue, anosmia, dysgeusia and cognitive problems. These can persist for months after infection, even in patients with only relatively mild initial (respiratory) symptoms (3, 4). There are millions of patients with long COVID worldwide. Yet our knowledge of the pathophysiology underlying these debilitating symptoms, as well as their long-term effects is still sorely lacking.

Several *post-mortem* histopathological studies have shown extensive inflammatory responses in organs of patients with acute COVID-19, including the brain (5, 6). Microglia activation is the most common pathology found in the brain of patients with COVID-19, with the highest levels of diffuse microglia activation in the olfactory bulbs, medulla oblongata, brainstem and cerebellum (5-7). However, to date there have been no *in-vivo* studies on neuroinflammation in patients with COVID-19 or after recovery. [^18^F]DPA-714 positron emission tomography (PET) provides a means of assessing neuroinflammation *in-vivo*. This is not only important because it may elucidate the pathophysiological cascade, but also and crucially because it may point to potential treatment approaches. Here, we report the results from the first two patients included in our [^18^F]DPA-714 PET study of microglia activation in long COVID.

## Patients and Methods

This study was approved by the Medical Ethics Review Committee of the Amsterdam UMC, location VU Medical Center, and registered under 2021-000781-15 in the European Union Drug Regulating Authorities Clinical Trials Database (EudraCT) and under NCT05371522 in ClinicalTrials.gov. After signing informed consent for this study, participants were thoroughly screened, provided a medical history, filled in several questionnaires addressing complaints and daily life functioning and participated in standardized neuropsychological assessment. Neuropsychological tests and questionnaires are specified in Table S1 and S2.

The first patient with long COVID included in our study was a Dutch woman, in her late fifties. She was healthy, worked full-time, and led a fulfilling life prior to developing a severe acute respiratory syndrome coronavirus 2 (SARS-CoV-2) infection. She was first infected in December 2020 (polymerase chain reaction (PCR)-test confirmed). During the acute phase of the infection, she was not hospitalized and did not need treatment.

However, since the infection, she has suffered from severe fatigue, concentration deficits, anosmia and parosmia, headaches, and some visual complaints. Her symptoms prevent her from working. Her medical history consisted of preexistent fibromyalgia (with subtle fatigue related complaints, that did not interfere with her work or daily functioning) and high cholesterol, which was at normal levels and stable with treatment. Her pre-existent fatigue-related complaints increased severely after infection with SARS-CoV-2. At the time of infection, she was not vaccinated against SARS-CoV-2. Four months following infection, she received her first dose of vaccine, and she was fully vaccinated (including a booster dose) one year after infection. Her numerous neurological symptoms started during the initial infection and her pre-existent fatigue worsened. Her symptoms remain present until this day, 15 months after infection.

The second patient with long COVID included was a Dutch male in his mid-sixties, who was healthy, fully functioning and worked full-time prior to the SARS-CoV-2 infection. He was infected in March 2020 (PCR-confirmed). During the acute phase of SARS-CoV-2 infection, he was hospitalized on a nursing ward for a total of 15 days. After ten days, he was admitted to the intensive care unit (ICU) for one night due to respiratory problems. During his hospitalization he received standard medication. Since the infection, he has suffered from severe fatigue and concentration deficits, and he was declared partially unfit for work for 75 %. He had no relevant past medical history. At the time of infection, he was not vaccinated against SARS-CoV-2. One year following infection, he received his first dose of vaccine, and he was fully vaccinated (including the booster dose) 21 months after infection. Although there has been some improvement, his symptoms remain present until this day, 24 months after infection.

### Genotyping

As the binding ability of the PET tracer [^18^F]DPA-714 depends on Translocator Protein (TSPO) genotype (8) with low, medium and high affinity binders, genotype of the RS6971 polymorphism in this gene was determined. Both patients were C/C, and therefore high affinity binders.

### Control subjects

To compare the level of neuroinflammation in long COVID patients to non-infected individuals, we reanalyzed historical data from a previous (published) [^18^F]DPA-714 study (9). This study was performed before 2019, ensuring that these subjects had not been infected with SARS-CoV-2. For (quantitative) comparison of the [^18^F]DPA-714 binding, we selected all three healthy control subject matching our patients with long COVID with respect to TSPO genotype (high-affinity binders). Control subject 1 was a female in her late fifties, control subject 2 a male in his late fifties, and control subject 3 a male in his late forties. For a comparison of [^18^F]DPA-714 metabolism, we included all available data from 7 healthy controls and 8 patients with MS.

### MRI

Long COVID patients underwent magnetic resonance imaging (MRI) at the Amsterdam UMC, location VUmc, on a 3.0T whole-body scanner (GE Signa, Discovery MR750). The scanning protocol included a high resolution 3DT1-weighted magnetization prepared rapid acquisition gradients-echo (MP-RAGE) image. Details are described elsewhere (9).

### PET

[^18^F]DPA-714 radiosynthesis was performed as described previously (10). Dynamic 60 minute [^18^F]DPA-714 PET scans with arterial blood sampling were acquired on the same Ingenuity TF PET-CT scanner (Philips Medical Systems, Best, The Netherlands) as described previously (9).

### Data analysis

To assess tracer metabolism, we compared the [^18^F]DPA-714 metabolites in the blood of the two long COVID patients to those of all other subjects available (9). We assessed activity concentration in whole blood, corrected for injected activity and tracer parent fraction (%), both from arterial blood samples.

Preprocessing of PET images has been described elsewhere (9). To quantify [^18^F]DPA-714 binding in whole brain gray matter we used a plasma input two tissue compartment model with blood volume parameter (2T4k_V_B_), as it was found previously to be the optimal model to quantify [^18^F]DPA-714 binding (9). Whole brain gray matter BP_ND_ values estimated using 2T4k_V_B_ were compared between two long COVID patients and three matching healthy control subjects. For visualization purposes we generated volume-of-distribution (V_T_) images using Logan plot analysis (11), using t* = 10 min, and divided those images by the whole brain grey matter k_1_/k_2_ ratio obtained by the plasma input 2T4k_V_B_ model, following a subtraction of 1. By doing so, Logan V_T_ images were corrected for the non-displaceable distribution volume resulting in BP_ND_ (=k_3_/k_4_) images for illustrative purposes. All quantitative whole brain grey matter BP_ND_ values reported are estimated using 2T4k_V_B_.

## Results

### Neuropsychological test and questionnaire scores

Neuropsychological test and questionnaire scores are provided in Table S1 and S2, respectively. Long COVID patient 1 had verbal memory deficits, mildly impaired sustained attention, presence of severe fatigue and concentration problems, and severe functional impairment. Long COVID patient 2 had visuo-constructive deficits, fluctuating sustained attention, presence of fatigue, severe concentration problems, and severe functional impairment.

### Whole blood activity concentration

We compared whole blood activity concentration of both long COVID patients to that of a previous [^18^F]DPA-714 cohort of healthy control subjects and patients with MS. As shown in Figure 1, tracer parent fraction and whole blood activity concentration corrected for injected activity for both long COVID patients was within the range of the retrospective data. This precludes that differences in tracer metabolism (and therefore availability) form an explanation for any differences in [^18^F]DPA-714 binding.

**Figure 1.**
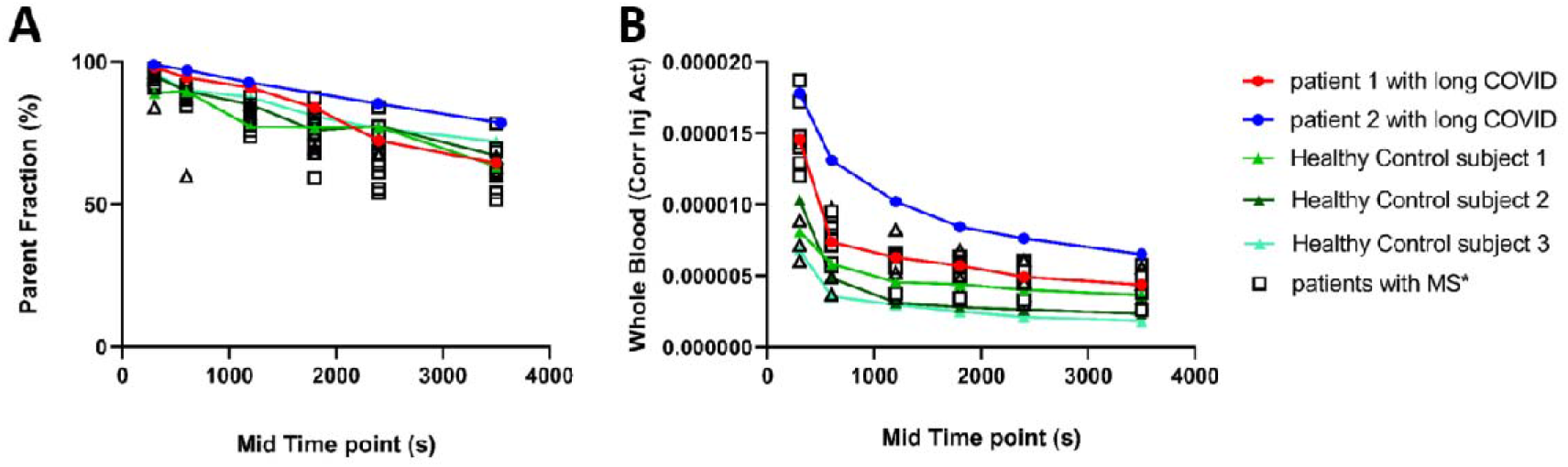
Whole blood activity concentration is in the range of a previous study for both long COVID patients. (**A**) Parent fraction and (**B**) Whole blood activity concentration corrected for injected activity (both parameters required to quantify tracer binding and thus determine the level of neuroinflammation) in two long COVID patients and the three healthy control subjects, plotted against data from the earlier [^18^F]DPA-714 cohort (of healthy control subjects and patients with multiple sclerosis (MS)).

### Imaging

Figure 2 shows the MRI and parametric BP_ND_ images of [^18^F]DPA-714 binding in the brain of the two long COVID patients and the three healthy control subjects. The MRI of the healthy control subjects and long COVID patient 1 were read as consistent with age by an experienced radiologist (FB), without relevant focal abnormalities. The MRI of long COVID patient 2 was also read as consistent with age, except for mild atrophy in the parietal region.

**Figure 2.**
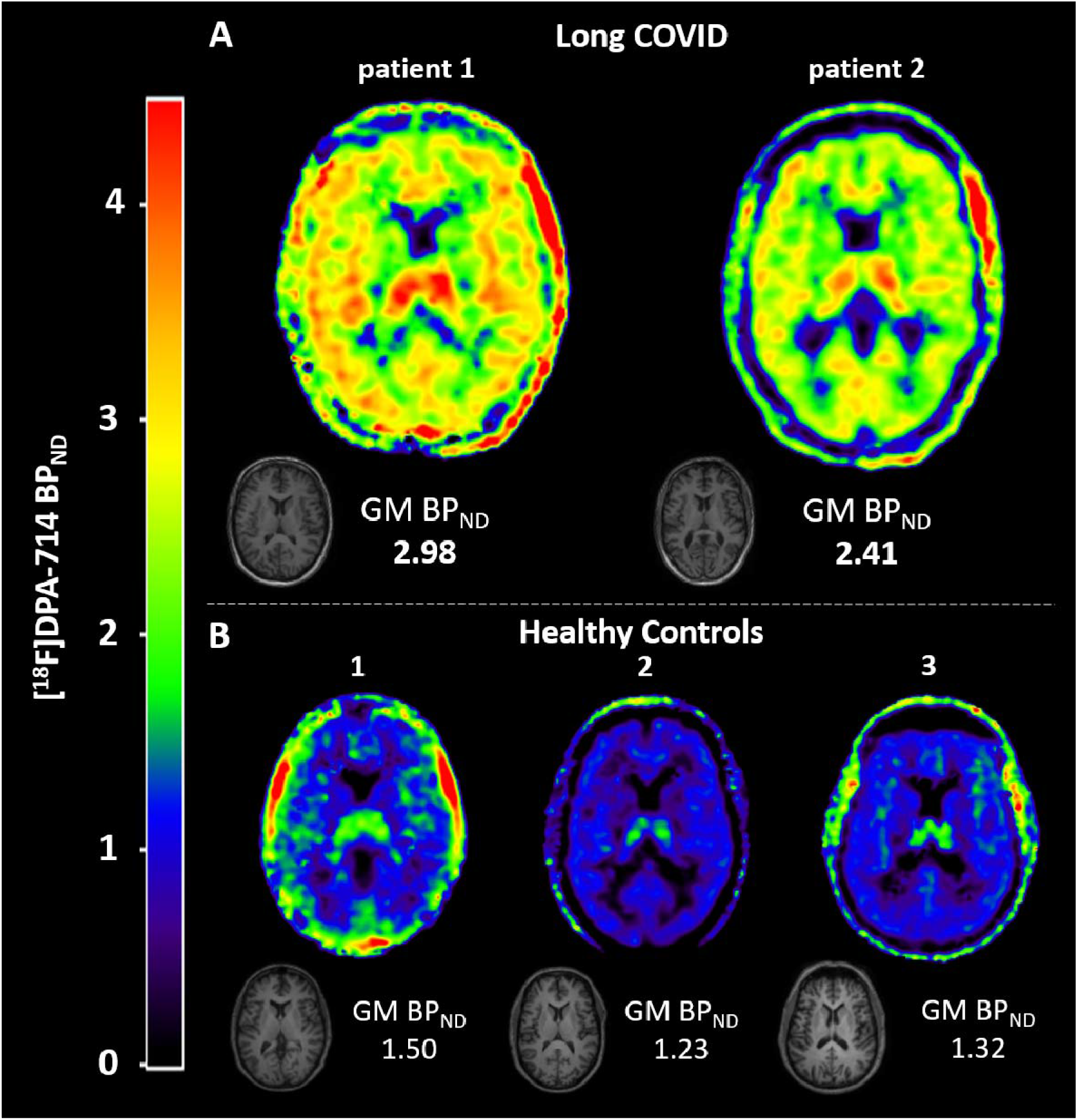
Neuroinflammation in two long COVID patients. To quantify [^18^F]DPA-714 binding in whole brain gray matter (GM) we used a plasma input two tissue compartment model with blood volume parameter (2T4k_V_B_). All quantitative whole brain grey matter binding potential (BP_ND_ (=k_3_/k_4_)) values reported are estimated using 2T4k_V_B_. For visualization purposes we generated volume-of-distribution (V_T_) images using Logan plot analysis (11), using t* = 10 min, and divided those images by the whole brain grey matter k_1_/k_2_ ratio obtained by the plasma input 2T4k_V_B_ model, following a subtraction of 1. By doing so, Logan V_T_ images were corrected for the non-displaceable distribution volume resulting in BP_ND_ (=k_3_/k_4_) images for illustrative purposes. (**A**) T1-weighted MRI and parametric images of [^18^F]DPA-714 binding in the brain of two long COVID patients. Higher binding potential (BP_ND_) values indicate more tracer binding and thus higher levels of neuroinflammation. Long COVID patient 1 showed severely elevated binding in all brain regions compared to the healthy control subjects. Long COVID patient 2 also showed elevated binding, with higher BP_ND_ values than the healthy control subjects. (**B**) T1-weighted MRI and parametric images of [^18^F]DPA-714 binding in the brain of three healthy control subjects.

As can be visually appreciated, long COVID patient 1 had severely elevated [^18^F]DPA-714 binding in all brain regions (Fig 2). Whole brain gray matter quantitative BP_ND_ (=k_3_/k_4_) obtained from the 2T4k_V_B_ model in long COVID patient 1 was increased by on average 121% relative to the healthy control subjects (Fig 2).

Long COVID patient 2 also had elevated [^18^F]DPA-714 binding (Fig 2), with an increase in whole brain gray matter BP_ND_ (=k_3_/k_4_) obtained from the 2T4k_V_B_ model by on average 79% relative to the healthy control subjects (Fig 2).

## Discussion

Long COVID is a worldwide problem, with high individual and societal cost, but no known pathophysiology. In this study, we report widespread and large increases in [^18^F]DPA-714 binding throughout the brain in the first two patients included in our *in-vivo* study of neuroinflammation. Although far from definitive, these findings are striking in extent and magnitude. As such, they implicate profound neuroinflammation in the pathophysiology of long COVID.

The extent of neuroinflammation in these patients with long COVID is remarkable. Whereas earlier *post-mortem* studies of acute COVID-19 patients have shown elevated neuroinflammation primarily in olfactory bulbs, medulla, brainstem and cerebellum, the current study suggests that the process of neuroinflammation may be more widespread in long COVID (5-7). Furthermore, as can be visually appreciated, we found high binding in the thalamus for both patients. This has also been reported in MS, using similar (TSPO) PET ligands (12). The thalamus is considered an important regulator, in relation to fatigue and cognitive functioning (13, 14) and may offer a clue towards the etiology of these symptoms in long COVID.

In addition to hinting at the pathophysiology of long COVID, neuroinflammatory processes could also be informative in relation to the prognosis of patients with long COVID. A recent study described cortical thinning and gray matter volume loss over time, as well as cognitive decline, in patients with long COVID (15). PET studies in MS have found that increased neuroinflammation is predictive of disease progression (16), suggesting a link between neuroinflammation and neurodegeneration on MRI. This may be relevant equally to long COVID. Furthermore, this raises the very important question of the ‘need to treat’ in the context of long COVID. The patients with long COVID in this study had widespread increases in neuroinflammation and substantial functional impairment, with only minimal abnormalities on MRI. Although these are preliminary findings and the exact relation between neuroinflammation, functional impairment and longer term structural brain changes is not yet established, taken together, these findings do raise the question whether treatment with anti-inflammatory drugs could be beneficial. Given the millions of people worldwide affected by this condition, the tremendous cost associated with it (to individuals and society) and the current, limited status of our knowledge, this is perhaps worthy of clinical investigation. Especially given recent findings from a study showing that vaccination prior to infection only seems to confer partial protection in the post-acute phase of the disease (17).

This study implicated neuroinflammation in persistent symptoms following COVID-19 infection and has several strengths. We were able to obtain spatial *in-vivo* information on neuroinflammation by using fully quantitative [^18^F]DPA-714 PET. We were able to compare the patients with three non-infected healthy control, with similar characteristics and scanned with the same methods on the same scanner, as well as blood data from a historical non-infected cohort. This permitted us to assess the severity of neuroinflammation. However, this study also has several limitations. First and foremost, we included only two patients with long COVID. These were the first two patients with long COVID included in this study, and we felt these data were too important not to publish at this stage. However, that does not negate that these remarkable findings will need to be replicated in a larger number of patients. Second, we had no comparable neuropsychological or questionnaire data available.

In conclusion, we report widespread and large increases in [^18^F]DPA-714 binding throughout the brain in the first two long COVID patients included in our *in-vivo* study. Long COVID represents a worldwide problem that comes at great cost to both individuals and society. Although our findings are far from definitive, they are striking in extent and magnitude. They implicate profound neuroinflammation in the pathophysiology of long COVID, and raise the question of whether (individualized) anti-inflammatory treatment could be beneficial.

## Data Availability

All data produced in the present study are available upon reasonable request to the authors

## Acknowledgements

We would like to thank the patients for their participation in the study, Prof. dr. S Durston for her critical appraisal of the manuscript, and Prof. dr. K Brinkman for his assistance with additional consent procedures.

