## Supplemental Material for "Long COVID is associated with extensive *in-vivo* neuroinflammation on [^18^F]DPA-714 PET"

**Table of content:**

Table S1: Neuropsychological test scores of the two patients with long COVID	Page 3

Table S2. Questionnaire scores of the two patients with long COVID	Page 3

**Table S1. Neuropsychological test scores of the two patients with long COVID.** Scores indicative of deficits are highlighted in bold.

|  | Score (normalized T-score) | |
| --- | --- | --- |
| Neuropsychological test | **Patient 1** | **Patient 2** |
| Symptom validity task | sufficient | sufficient |
| The Montreal Cognitive Assessment (MOCA)(25) | 26/30 | 27/30 |
| Memory |  |  |
| Dutch version of the Rey Auditory Verbal Learning Test (RAVLT) – immediate recall | **30 (T25)** | 55 (T62) |
| Dutch version of the Rey Auditory Verbal Learning Test (RAVLT) – delayed recall | **5 (T29) (T46 when corrected for immediate recall)** | 11 (T57) (T44 when corrected for immediate recall) |
| Dutch version of the Rey Auditory Verbal Learning Test (RAVLT) – recognition | **27** | 30 |
| D2 |  |  |
| F% – percentage errors | 7 (T41) | 12 (T36) |
| Tn-F – total processed items (error corrected) | 303 (T40) | 430 (T60) |
| CP – concentration prestation | 116 (T40) | 145 (T52) |
| VT – variation in speed | 15 (T42) | 19 (T35) |
| Attention/Executive functioning |  |  |
| Trail Making Test – version A | 23 (T60) | 20 (T68) |
| Trail Making Test – version B | 71 (T47) | 71 (52) |
| TMT B/A | 3.1 (T42) | 3.6 (T43) |
| Digit Span Forward | 12 (T45) | 16 (T62) |
| Digit Span Backward | 7 (T41) | 14 (T73) |
| Stroop word test | 36 (T58) | 36 (T59) |
| Stroop color test | 42 (T66) | 50 (T57) |
| Stroop color-word test | 78 (T54) (T45 when corrected for Stroop color test) | 96 (T48) (T44 when corrected for Stroop color test) |
| Visuospatial functioning |  |  |
| Rey Complex Figure – copy | 36 (100^th^ percentile) | **27 (<10^th^ percentile)** |
| Rey Complex Figure – delayed recall | 23 (55^th^ percentile) | **12 (<10^th^ percentile)** |
| Language |  |  |
| Letter Fluency (K-O-M) | 40 (T49) | 57 (T66) |
| Letter Fluency category animals | 25 (T52) | 24 (T50) |

**Table S2. Questionnaire scores of the two patients with long COVID.** Cut-off for severe fatigue on the CIS-fatigue is ≥35. Cut-off for severe concentration problems on the CIS-concentration is ≥15. Cut-off for severe impairment on daily functioning on the Work and Social Adjustment Scale is ≥10.

|  | Score/Total | |
| --- | --- | --- |
| Questionnaire | **Patient 1** | **Patient 2** |
| Fatigue subscale of the Checklist Individual Strength (CIS-fatigue) | **53/56** | 32/56 |
| Concentration subscale of the Checklist Individual Strength (CIS-concentration) | **18/35** | **19/35** |
| Beck Depression Inventory for Primary Care (BDI-PC) | 1/21 | 0/21 |
| Generalized Anxiety Disorder – 7 (GAD-7) | 2/21 | 0/21 |
| Work and Social Adjustment Scale | **21/40** | **16/40** |
